# SARS-CoV-2 seroprevalence among Vancouver public school staff in British Columbia, Canada

**DOI:** 10.1101/2021.06.16.21258861

**Authors:** David M. Goldfarb, Louise C. Mâsse, Allison W. Watts, Sarah M. Hutchison, Lauren Muttucomaroe, Else S. Bosman, Vilte E. Barakauskas, Alexandra Choi, Michael A. Irvine, Frederic Reicherz, Daniel Coombs, Collette O’Reilly, Sadaf Sediqi, Hamid R. Razzaghian, Manish Sadarangani, Sheila F. O’Brien, Pascal M. Lavoie

**Affiliations:** BC Children’s Hospital Research Institute, University of British Columbia, Vancouver, BC, Canada; Department of Pathology and Laboratory Medicine, University of British Columbia, Vancouver, BC, Canada; BC Children’s and Women’s Health Centre, Vancouver, BC, Canada; School of Population and Public Health, University of British Columbia, Vancouver, BC, Canada; Department of Pediatrics, Faculty of Medicine, University of British Columbia, Vancouver, BC, Canada; Vancouver Coastal Health, Office of the Chief Medical Health Officer, Vancouver, BC, Canada; British Columbia Centre for Disease Control, Vancouver, BC, Canada; Department of Mathematics and Institute of Applied Mathematics, University of British Columbia, Vancouver, BC, Canada; Vancouver School District, Vancouver, BC, Canada; Vaccine Evaluation Center, BC Children’s Hospital Research Institute, Department of Pediatrics, University of British Columbia, Vancouver, BC, Canada; Epidemiology & Surveillance, Canadian Blood Services, Ottawa, Ontario, Canada; School of Epidemiology & Public Health, University of Ottawa, Ottawa, Ontario, Canada

**Keywords:** SARS-CoV-2, COVID-19 seroprevalence, schools, occupational risk

## Abstract

**Importance:** Contact-tracing studies suggest minimal secondary transmission in schools. However, there are limited school data accounting for asymptomatic cases, particularly late in the 2020/21 school year, and in the context of uninterrupted in-person schooling and widespread community transmission.

**Objectives:** To determine the SARS-CoV-2 seroprevalence in a sample of school staff, compared to the community, and to COVID-19 rates among all students and staff within the same school population.

**Design:** Incident COVID-19 cases among students and school staff using public health data, with an embedded cross-sectional serosurvey among school staff sampled from February 10 to May 15, 2021, comparing to age, sex and geographic location-matched blood donors sampled in January 2021.

**Setting:** Vancouver School District (British Columbia, Canada) from kindergarten to grade 12.

**Participants:** Active **s**chool staff enrolled from February 3 to April 23, 2021.

**Main outcome measures:** SARS-CoV-2 antibodies in a sample of school staff using spike (S)-based testing (unvaccinated staff) or N-based serology testing (vaccinated staff).

**Results:** The incidence of COVID-19 cases among students attending in-person was 9.8 per 1,000 students during the 2020/21 school year (N = 47,280 students), and among staff was 13 per 1,000 since the beginning of the pandemic (N = 7,071 active school staff). In total, 1,689 school staff (64% elementary, 28% secondary, 8.3% school board staff or multiple grades) completed the questionnaire, 78.2% had classroom responsibilities, and spent a median of 17.6 hours in class per week [IQR: 5.0 – 25 hours]. Although 21.5% (363/1,686) reported close contact with a COVID-19 case, only 1.4% (24/1688) of the school staff reported having had a positive viral nucleic acid test. Of this group, five believed they acquired the infection at school. The adjusted seroprevalence in staff who gave blood (1,556/1,689, 92.1%) was 2.3% [95%CI: 1.6 – 3.2%] compared to 2.3% [95%CI: 1.7 – 3.0%] in blood donors.

**Conclusion and relevance:** Despite high reported COVID-19 cases among students and staff, and frequent within-school exposures, we found no detectable increase in seroprevalence among school staff above the community seroprevalence. These findings corroborate claims that, with appropriate mitigation strategies, in-person schooling is not associated with significantly increased risk for school staff.

**Key Points:** *Question:* What was the prevalence of COVID-19 infections in school staff who maintained in-person schooling during the 2020/21 school year in Vancouver, British Columbia, and how does it compare to the risk of COVID-19 infection in the community.

*Findings:* As of March 4, 2021, the incidence of COVID-19 cases among school staff was 13 per 1,000 (N = 7,071 school staff) since the beginning of the pandemic. In a cross-sectional seroprevalence analysis from February 10 to May 15, 2021, the adjusted seroprevalence among a sample of school staff (N = 1,556) was 2.3% [95%CI: 1.6 – 3.2%], compared to 2.3% [95%CI: 1.7 – 3.0%] in 1:2 age, sex and geographical location (by postal code)-matched reference group of blood donors.

*Meaning:* We found no detectable increase in seroprevalence among school staff above the community seroprevalence. These findings corroborate claims that, with appropriate mitigation strategies in place, in-person schooling is not associated with significantly higher risk for school staff.

## Introduction

SARS-CoV-2 forced over a billion students out-of-school globally in the Spring 2020. Decisions to close schools, motivated by high case mortality in populations, had serious implications for children’s emotional, social, physical and educational outcomes^1^. The risk of secondary SARS-CoV-2 transmission within schools has been heavily debated. On one hand, viral culture studies suggest that children may be less infectious than adults^2^, contact tracing studies show low rates of in-school transmission^3-13^, and surveillance studies demonstrate little increased transmission when schools re-opened^14-19^. On the other hand, seroprevalence studies have been conducted to account for asymptomatic transmission, but many studies have reported data early in the pandemic, or in the setting of partial school closure^15,20-22^.

In the spring of 2020, Public Health authorities in British Columbia (BC) ordered a cessation of in-person schooling provincially, with a transition to remote learning from home. Like most of the world, the province went under a nearly complete lockdown between March and early June when most sectors of the economy were paused. While BC reported relatively low community viral transmission early in the first pandemic wave, roughly >50 times more COVID-19 cases were reported (121,762 cases reported in a population of 5,017,000) between July 1, 2020 and April 23, 2021, compared to February to May 2020.

Despite increasing cases in late summer, BC was unique within Canada in that it maintained in-person schooling for the entire duration of the 2020/2021 school year starting September 8, 2020, except for winter (December 18, 2020 to January 4, 2021) and spring (March 12 to March 29, 2021) breaks.

The main goal of this study was to determine the SARS-CoV-2 seroprevalence in school staff in Vancouver public schools during the 2020/21 school year. The secondary objectives were to compare the seroprevalence in school staff to a reference population of matched Canadian blood donors, and to report on the incidence of COVID-19 cases among all students and school staff. We hypothesized that with mitigation measures the occupational risk of SARS-CoV-2 infection among school staff would remain low and comparable to the community risk.

## Methods

### Study design

The baseline questionnaire and serology data collected among active school staff of the Vancouver School District (the District) who are being followed for one year, was used to determine SARS-CoV-2 seroprevalence. Seroprevalence in school staff was compared to an age, sex and geographic location-matched sampled of blood donors collected in January 2021. In addition, we obtained retrospective data on COVID-19 cases in all staff and students attending the District from Vancouver Coastal Health (VCH, detailed below).

### Participants

*School staff* self-enrolled from February 3 to April 23, 2021 after receiving an introduction email from school principals from the District in early February 2021, inviting them to register online at: https://www.bcchr.ca/COVIDatschools, for both a questionnaire and to provide blood for serology testing. A flyer was posted on the District website, and reminder emails were also sent. Interested participants completed a screener to identify whether they met eligibility criteria. Staff were included if they were a current, full or part-time staff member (confirmed by District email address). Staff who reported being temporary staff, on-leave, or on-call with no reported classroom time were excluded. Informed consent was obtained from all school staff. The study was approved by the University of British Columbia Children’s and Women’s Research Ethics Board (H20-03593).

*Blood donors* were screened prior to donation to ensure they were in good health, including questions about COVID-19. People were ineligible to donate blood if they had a recent COVID-19 infection two weeks after symptoms resolved, or were hospitalized within 3 weeks before.

### Study setting

The District is a large, urban school district with 89 elementary schools and 18 secondary schools (47,280 students and 7,071 school staff) located in the city of Vancouver (BC, Canada ∽600,000 population in the city of Vancouver with 2.6 million population in the urban area). Following a complete closure in March 2020, schools opened in a limited fashion, except for students who use English as a second language and those with complex learning needs who were able to attend in-person 5 days/week until June 30, 2020. On September 8, 2020 schools reopened for the 2020/21 school year, except for a winter break from December 18, 2020 to January 4, 2021, and spring break from March 12 to 29, 2021. COVID-19 mitigations measures implemented in District schools as well as indications for viral testing are detailed in **Appendix 1**.

### Data Collection

Data was collected from school staff using a questionnaire that asked, among others, about risk factors for COVID-19, household structure, physical distancing behavior, close contacts with COVID-19 cases (defined by asking: *“someone diagnosed with COVID-19 with whom you’d been within two meters of for greater than two minutes”*), history of viral testing (including dates and symptoms) and vaccination, etc.^23^ A second questionnaire about mental health and vaccine perception was also administered but not reported in this paper. COVID-19 vaccination in blood donors was collected by asking at the time of blood donation.

To estimate the degree of exposure to known COVID-19 cases, we obtained data from VCH’s Case and Contact Management Interface. The District provided student and staff lists to VCH, which linked the data to determine the incidence of SARS-CoV-2 infection among students and staff in District schools. Adult education staff were excluded. Staff and students affiliated with Vancouver Alternate Secondary School programs were counted as attending a single school for the purposes of incidence calculations.

Given that the history of viral testing in the prospective school staff sample was obtained via questionnaires, we selected the median date of questionnaire completion (March 4, 2021) as the end date for data extraction. We extracted all lab-confirmed, probable, and epidemiologically-linked COVID-19 cases reported to VCH. To assess the incidence of known infection among staff over the course of the pandemic, we calculated the incidence of reported staff cases from January 15, 2020 (corresponding to the first case reported to VCH) to March 4, 2021. Similarly, exposure to student cases during the school year was estimated by calculating the incidence from September 8, 2020 to March 4, 2021.

### Serology testing

Blood samples were collected from February 10 to May 15 2021, at clinics set-up in various participating Vancouver schools, at the BC Children’s Hospital or outpatient clinical laboratories in metro Vancouver. The presence of antibodies against SARS-CoV-2 was used as a marker of prior COVID-19 infection, using dual S- and N-based serology testing, where S-based serology was used in unvaccinated participants and N-based serology testing was used with vaccinated participants (**Supplemental Figure 1**).

Antibodies directed against the spike (S1) protein were detected using the Ortho T *VITROS™* Anti-SARS-CoV-2 *Total antibody assay* (Ortho IgG; Ortho Clinical Diagnostics, Rochester, NY), a Health Canada and FDA-licensed qualitative assay which detects all types of antibodies (IgA, IgG and IgM). S-based serology testing was done on a Vitros 5600 analyser at the BC Children’s & Women’s Hospital Laboratory, which is accredited for clinical testing. Literature and in-house validation demonstrated this assay can identify both symptomatic and asymptomatic infected individuals >7 days post illness onset with a sensitivity between 90.7% and 97.7%, and specificities between 99.4% and 100% ^24,25^. Specimens were considered reactive at a cut-off index ≥1.00. All S-tested negative samples with S-antibody indexes

>99^th^ centile were also confirmed to be negative on the Roche assay. Testing for anti-nucleocapsid (N) protein SARS-CoV-2 antibodies was performed using the Roche Elecsys*™* Anti-SARS-CoV-2 (Roche T; Roche, USA). This qualitative total antibody assay is Health Canada and FDA-licensed with reported sensitivity of 88.5% – 100% at least 14 days post-COVID-19 onset and specificity of 99.8% –100% ^26-28^. Testing was performed on a Cobas e601 analyzer at St. Paul’s Hospital Laboratory.

Blood donors were tested for both SARS-CoV-2 spike and N antibodies using the Roche Elecsys™ Anti-SARS-CoV-2 S and Anti-SARS-CoV-2 (Roche, USA) assays, respectively, on a Cobas e801 analyzer, and assigned using a similar S/N strategy for vaccinated / unvaccinated cases (**Supplemental Figure 1**).

### Statistical analyses

Blood donors were identified to match study participants by age and sex ± 2 years and two donors randomly selected without replacement for each study participant. As not all participants could be matched, criteria were relaxed to include additional donors (**Supplemental Data**). The Rogan-Gladen estimator was used to calculate the true prevalence adjusting for test specificity and sensitivity, with 95% confidence intervals estimated using Blaker’s method.^29^ For seroprevalence in school staff and blood donors, sensitivities of 95.3% and 96.6%, and specificities of 100% were used for the two S-based assays, respectively.^25,30^

## Results

### COVID-19 cases from VCH contact tracing data in District schools (N = 47280 students and N = 7071 school staff)

During the 2020-2021 school year 46879 students attended District schools in-person, and 401 students attended an Alternate District School (total of 47280 students). As shown in **Figure 1**, the overall weekly rates of reported COVID-19 cases among staff and students during the pandemic followed a trend similar to the weekly rates among Vancouver residents.

The population-level incidence of COVID-19 cases among students (including Vancouver Alternate Secondary Schools) during the 2020/21 school year was 9.8 cases per 1000 students, ranging from 0 to 63 cases per 1000 between schools (**Supplemental Figure 2**). Among schools with at least 1 student case, the median number of student cases was 3, and the median school population was 376 students.

In addition, 67 out of the 107 schools (62.6%) had no staff members diagnosed as confirmed, probable, or epidemiologically-linked COVID-19 cases since the beginning of the pandemic. Twenty-six of the 40 schools (65%) with staff cases had only one staff case. Including staff of Vancouver Alternate Secondary Schools, the incidence of reported cases from January 15, 2020 to March 4, 2021 among staff within specific schools ranged from 0 cases per 1000 staff to a maximum of 167 COVID-19 cases per 1000 staff. Among schools with at least 1 staff case, the median number of staff cases was 1, and the median size of each school’s staffing complement was 46.5 staff members. All schools with incidences higher than 80 COVID-19 cases per 1000 staff had only 1 or 2 staff cases among a staffing complement of under 25 staff. The incidence of reported COVID-19 cases from January 15, 2020 to March 4, 2021 was 13 cases per 1000 classroom staff, and 14 cases per 1,000 non-classroom staff (**Supplemental Table 1**).

### COVID-19 cases self-reported in school staff sample (N = 1689)

Staff COVID-19 cases reported to VCH were compared to questionnaire data. In total, there were 2162 access to the screener, of which 1743 staff identified themselves, provided contact information and consented on-line (**Figure 2**). The characteristics of 1689 staff who completed the questionnaire, corresponding to 23.9% of eligible staff (**Table 1**).

**Table 1:** Baseline characteristics of school staff sample.

| Variable | n<sup>#</sup> | Completed questionnaire (n=1689) | n<sup>#</sup> | Completed serology testing (n=1556) |
| --- | --- | --- | --- | --- |
| Age (mean ± SD) | 1684 | 45.4 ± 10.4 | 1556 | 45.7 ± 10.3 |
| Sex, % female, n (%) | 1681 | 1355 (80.6%) | 1550 | 1257 (81.1%) |
| Canadians of indigenous origin, n (%) | 1688 | 31 (1.8%) | 1555 | 31 (2.0%) |
| Ethnicity, n (%) | 1689 |  | 1556 |  |
| White, Caucasian |  | 1175 (69.6%) |  | 1084 (69.7%) |
| South Asian |  | 65 (3.9%) |  | 57 (3.7%) |
| Chinese |  | 277 (16.4%) |  | 257 (16.5%) |
| Black |  | 12 (0.7%) |  | 12 (0.8%) |
| Filipino |  | 35 (2.1%) |  | 33 (2.1%) |
| Latin American |  | 26 (1.5%) |  | 26 (1.7%) |
| Arab |  | 4 (0.2%) |  | 3 (0.2%) |
| Southeast Asian |  | 32 (1.9%) |  | 27 (1.7%) |
| West Asian |  | 1 (0.1%) |  | 1 (0.1%) |
| Korean |  | 11 (0.7%) |  | 9 (0.6%) |
| Japanese |  | 39 (2.3%) |  | 36 (2.3%) |
| Other/no answer |  | 62 (3.7%) |  | 57 (3.7%) |
| Classroom workers*, n (%) | 1688 | 1320 (78.2%) | 1555 | 1212 (77.9%) |
| Contact time with students (hrs/wk, median [IQR]) | 1684 | 17.6 [5.0-25.0] | 1552 | 17.5 [4.6-25.0] |
| School level, n (%) | 1689 |  | 1556 |  |
| Elementary |  | 1076 (63.7%) |  | 992 (63.8%) |
| Secondary |  | 474 (28.1%) |  | 436 (28.0%) |
| Work at multiple levels |  | 55 (3.3%) |  | 48 (3.1%) |
| School district office only |  | 84 (5.0%) |  | 80 (5.1%) |
| No. people living in household (median [IQR]) | 1685 | 3 [2-4] | 1552 | 3 [2-4] |
| No. people in household is essential worker (median [IQR]) | 1671 | 0 [0-1] | 1541 | 0 [0-1] |
| At least one co-morbidity<sup>†</sup>, n (%) | 1689 | 409 (24.2%) | 1556 | 379 (24.4%) |
| Smoker, n (%) | 1686 | 46 (2.7%) | 1553 | 41 (2.6%) |
| Travelled outside BC since Jan '20, n (%) | 1687 | 278 (16.5%) | 1554 | 252 (16.2%) |

<sup>#</sup>N with data available.
\* Those who reported being a Teacher, Teacher Librarian, Resource Teacher, Student Support Worker, or Family and Youth Worker in response to the question: *What is your job title? Teacher/Teacher Librarian/Resource Teacher/Student Support Worker/Family and Youth Worker/Administrator (Principal, Vice Principal)/Administrative Assistant/Maintenance Staff/School Board Office Staff/Other.*
<sup>†</sup>any the following: hypertension, diabetes, asthma, chronic lung disease, chronic heart disease, chronic kidney disease, liver disease, cancer, chronic blood disorder, immunosuppressed, chronic neurological disorder.

Notably, 63.7% of study participants were elementary school staff and 28.1% were secondary school staff (**Table 1**), which align with District data (not shown). Overall, a majority (78.2%; n = 1320) of school staff were classroom staff, and spent a median of 17.6 hours of contact time with students per week (**Table 1**). The District estimated that 5091 staff have classroom responsibilities. Therefore, we estimate that the study enrolled at least ∼26% of all staff with classroom responsibilities.

About one third (37%) of school staff lived with an essential worker, predominantly in the social services, education/research/healthcare, construction, maintenance and skilled trades, and food sectors (**Table 2**).

**Table 2:**
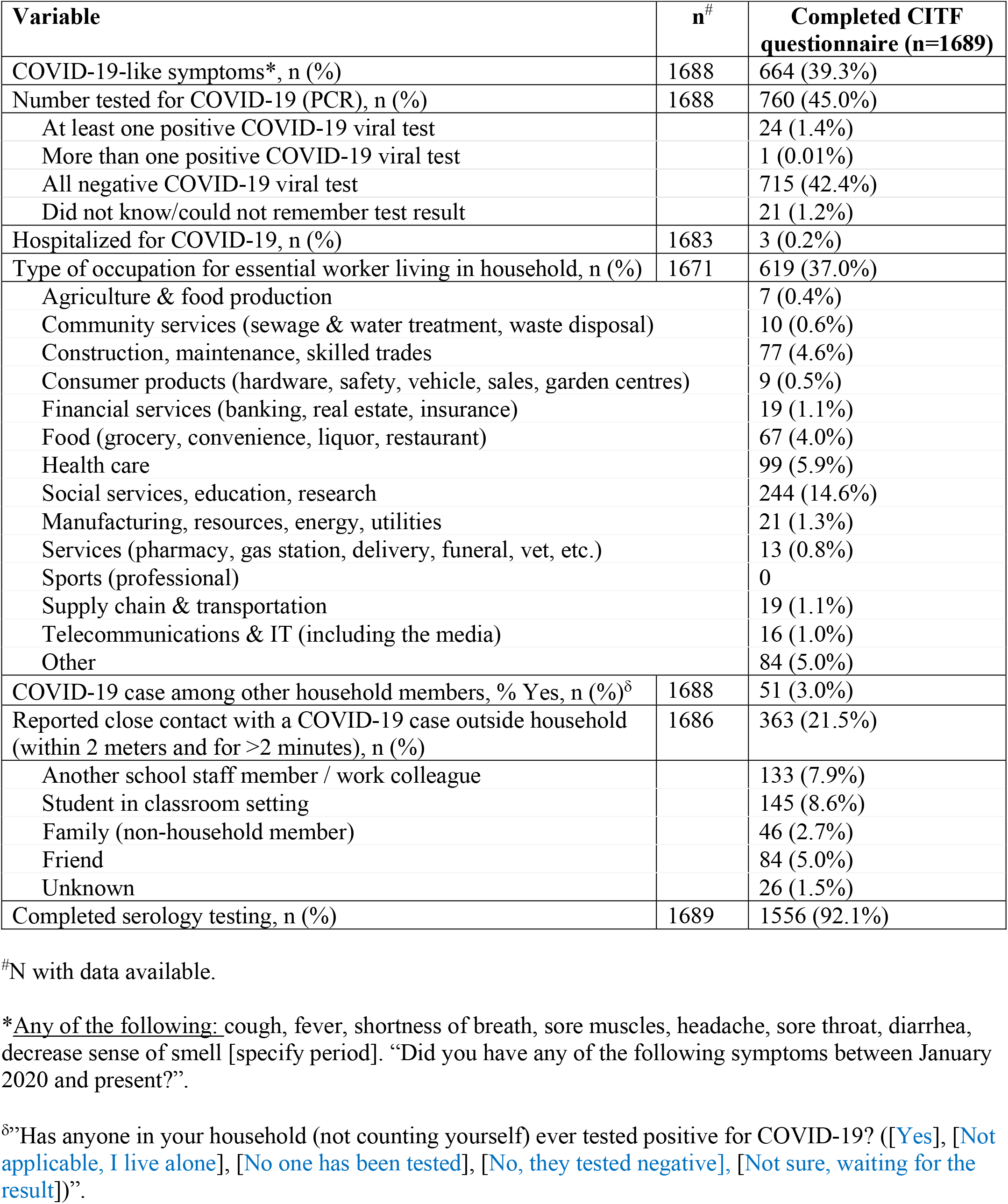
Reported COVID-19 exposures and PCR outcomes among school staff.

Among the school staff who completed the questionnaire, 51 reported a positive viral test among their household members (**Table 2**). In total, 363 (21.5%) reported a history of close contact with a COVID-19 case at or outside school, but only 24 reported ever testing positive by nucleic acid testing (**Table 2**). Four (16.7%) tested positive by nucleic acid testing prior to the beginning of classes in September 2020.

In total, 24 of 1689 school staff self-reported a positive viral test which represents an incidence rate of COVID-19 case of 1.4%. Of the 24 school staff who reported a positive viral test, 5 (21%) reported close contact with a student or co-worker case, including one who required hospitalization during the 2020/21 school year. Seven (29%) reported close contact with a friend or family member with COVID-19, and one reported close contact with both a co-worker and family member with COVID-19. Eleven had unknown sources of acquisition and were not aware of any close contact with a COVID-19 case.

### SARS-CoV-2 seroprevalence in school staff (N = 1556)

Of 1689 school staff with prospective questionnaire data, 1556 completed serology testing (median date: March 11, 2021). Serology results for vaccinated and SARS-CoV-2-infected staff are shown in **Supplemental Table 2** and **3**, respectively. In total, 35 tested positive for SARS-CoV-2 by serology. Therefore, the unadjusted prevalence was 2.2% (95%CI: 1.6 – 3.1%), and the seroprevalence after adjusting for the sensitivity and specificity of the test was 2.3% (95%CI: 1.6 to 3.2%). Of the 35 school staff who seroconverted, 29 worked in a classroom setting and one did not work in a classroom setting, but reported more than 20 hours of contact time with students per week, for a seroprevalence also of 2.3% (95%CI: 1.5 – 3.1%). The proportion of staff who tested positive for SARS-CoV-2 by serology between secondary and elementary schools (**Table 3**) corresponded to the proportion of staff in each school level (**Table 1**). In comparison, the unadjusted seroprevalence in age, sex and geography-matched blood donors was 2.0% (95%CI: 1.5 – 2.7%), and 2.3% (95%CI: 1.7 – 3.0%) after adjusting for the sensitivity and specificity of the serology test.

**Table 3:** Seropositive cases according to school education level where school staff teaches/assists.

| **School** | **Frequency** | **Percent cases** |
| --- | --- | --- |
| Elementary | 19 | 54.3 |
| Secondary | 9 | 25.7 |
| Multiple / mixed | 3 | 8.57 |
| School board office | 4 | 11.4 |

## Discussion

To the best of our knowledge, this study is one of the largest to report seroprevalence estimates in the school setting in the later phases of the pandemic in the context of in-person schooling and widespread viral transmission. This study found that the seroprevalence among staff in Vancouver public schools was low after a period of widespread community transmission. Results were consistent with both self-reported infection as well as COVID-19 cases reported by VCH. Findings are in keeping with modelling studies ^31,32^ and data from the UK where low seroprevalence was also measured in teachers, but this was earlier in the pandemic^15^. Moreover, despite that the seroprevalence in this study represents an approximately three-fold increase relative to previous estimates of 0.55% to 0.6% obtained from Vancouver residents in spring 2020^33-35^, it remained comparable to age/sex and area of residency-matched blood donors.

A major advantage of the current study is that it was conducted in BC, one of the few, if not only (as far as we are aware) jurisdictions in North America that maintained in-person schooling during the 2020/21 school year. Study results are drawn from a large sample of staff, including a majority of those exposed to COVID-19 in the classroom. The use of S-based serology assays identified COVID-19 cases up to a year before. The study utilized sensitive serology testing to identify cumulative SARS-CoV-2 cases that may have not come to clinical attention, but could still contribute to the transmission chain^36^. Conversely, the N-based serology test allowed us to account for vaccinated staff towards the end of recruitment.

Among our study participants, 21.5% (363) of school staff reported a close contact with a COVID-19 case, and the majority (76.6%, 278/363) identified contact with a COVID-19 case at school. These data alone could reinforce the perception that schools are a risky environment. However, we could not find evidence to substantiate the perception that a large number of asymptomatic infections have been missed through contact tracing, and thus we were able to provide a more accurate depiction of viral transmission. Despite the high frequency of school staff who reported symptoms (**Table 2**), 90.1% (598/664) had no serological evidence of infection using a conservative testing strategy. Under-ascertainment by viral testing could have been related to the use of targeted testing up until April 2020. However, the relatively high proportion (60%) of cases diagnosed by nucleic acid testing who tested positive via serology suggests good access to viral testing in this specific testing, during the study period.

Mitigation strategies employed in BC schools have been shown elsewhere to minimize risk in educators to a level comparable to the risk in the community ^37,38^. Although non-medical masks were encouraged, but not required in schools until February 2021 (grades 8-12) and end of March 2021 (grades 4-12) (**Appendix**), and are still not required for K-3 students – a situation that is unique in Canada - we also did not detect any meaningful difference in seroprevalence between elementary and secondary school staff.

This study has limitations. First, study volunteers are typically healthier, raising a possibility that seroprevalence estimates were underestimated. To estimate this bias, the incidence of COVID-19 cases based on self-report (1.4%) was compared to the contact tracing data in classroom staff across the entire District (1.3%) and suggests that we did not under sampled those who are in direct contact with students. Second, the seroprevalence of school staff was compared to matched blood donors which may underestimate the community prevalence as the blood donors serology were taken in January 2021, prior to sampling of the school cohort, which could only reinforce our conclusions. As others have found, serologic testing in blood donors in general does reflect overall seroprevalence in the community^39-42^ and we would expect the seroprevalence in the reference group of blood donors for the same period to be higher, which reinforces our study conclusion. Consistent with this, seroprevalence estimates based on anonymized, residual specimens collected by the BC Centre for Disease Control in January 2021 from working age adults (attending one of ∼80 diagnostic service centres in the only outpatient laboratory network in the greater Vancouver area) ranged from 3-4% (DM Skowronski, personal communication). Second, the study had limited power to detect small differences between seroprevalence in school staff and the community. However, it is unlikely that small differences would drastically change the public health recommendations that can be made from the data. Third, exposure risks may differ between school and community settings, which needs to be taken into account when attempting to generalize our findings.

In conclusion, this study shows no detectable increase in SARS-CoV-2 infections in school staff working in Vancouver public schools following a period of widespread community transmission (October – May 2021). The combination of population-level COVID-19 data based on nucleic acid testing in symptomatic cases and sensitive serology testing among school staff provide strong evidence that their risk of COVID-19, in the context of mitigation strategies, is not substantially higher than the community.

## Data Availability

The data will eventually be made publicly available through the COVID-19 Immunity Task Force

## Authors’ contributions

LCM and PML obtained funding as Co-PIs; DMG, AW, MAI, DC, PML and LCM designed the original study concept; SH was involved in the earliest stages of the study, helping with funding applications, study design, ethics applications and data interpretation; MS also advised initial study design; FR helped review the literature; EB constructed and managed the data collection database with support from LCM and AW; LM set-up and coordinated the recruitment of participants; SS and HRR processed all blood samples; VEB supervised the collection of blood samples; AC supervised the analysis of VCH case data among students and school staff; MAI advised statistical study design and analyzed the main seroprevalence estimates; SO provided and analysed matched data from Canadian blood donors; AW performed all other data analyses; CO facilitated communications within the District during the study; DMG & PML drafted the first manuscript with specific sections written by AW, SH, VEB, MAI, AC, CO and LCM. All authors revised the manuscript and approved its final version.

## Acknowledgements

We first of all would like to thank the teachers and all other Vancouver School District staff who participated in the study and have been working tirelessly during this pandemic, and the District leadership, particularly Suzanne Hoffman and David Nelson for providing full support during this study; Kathy O’Sullivan for ongoing work reviewing District communications, documents and liaising with school partners throughout the study; Esther Alonso-Prieto for help with the human resource and financial management of this study; Brandon Bates, John Bhullar and Chemistry Laboratory staff at Children’s and Women’s Hospitals and the BC Children’s Hospital Biobank staff for help with sample collection and processing; the District and BC Children’s Hospital communication teams, Kim Schmidt and Lea Separovic for help with advertising; Laura Burns and Janet Simons, from the St-Paul’s Hospital Laboratory, for coordinating the Roche N-based serology testing; Nalin Dhillon from VCH for his analytic expertise; VCH’s Office of the Chief Medical Health Officer, the BC Centre for Disease Control, Dr. Sarka Lisonkova and KS Joseph for ongoing discussions that guided this study, and Steven Drews and Qi-Long Yi for consultation and assistance with matched donor data from Canadian Blood Services.

---

**Supplemental Figure 1:**
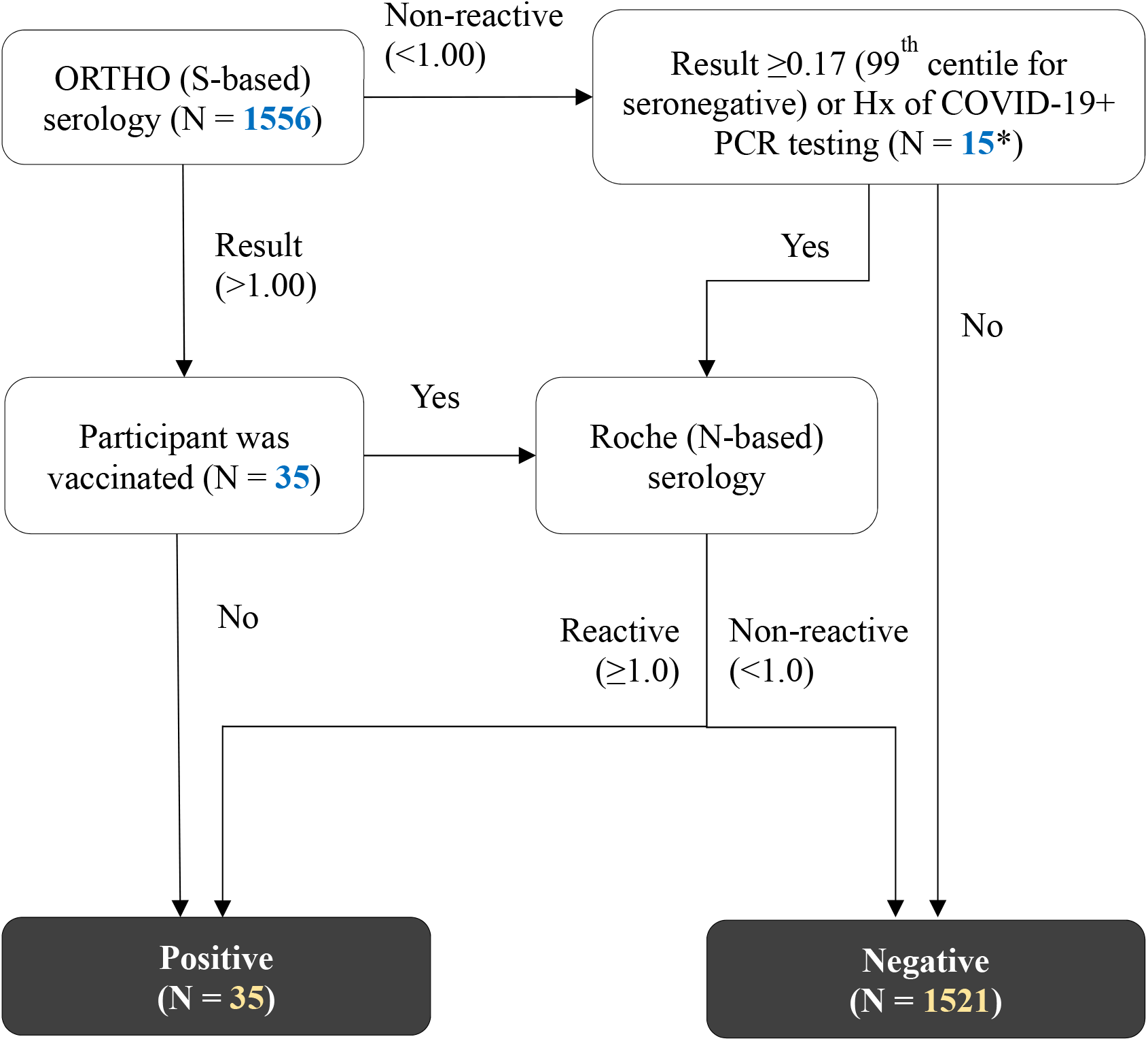
Seroprevalence case assignment strategy. To distinguish between antibodies due to COVID-19 versus antibody responses to vaccination, a dual, stepwise serology testing strategy was employed, where more sensitive S-based testing (using ORTHO assay) was used in unvaccinated school staff who composed the majority of our study sample, and virus-specific N-based antibody testing (using Roche assay) was used in vaccinated school staff. *One case reported PCR positive testing had subthreshold S-based serology reactivity of 0.39.

**Supplemental Figure 2:**
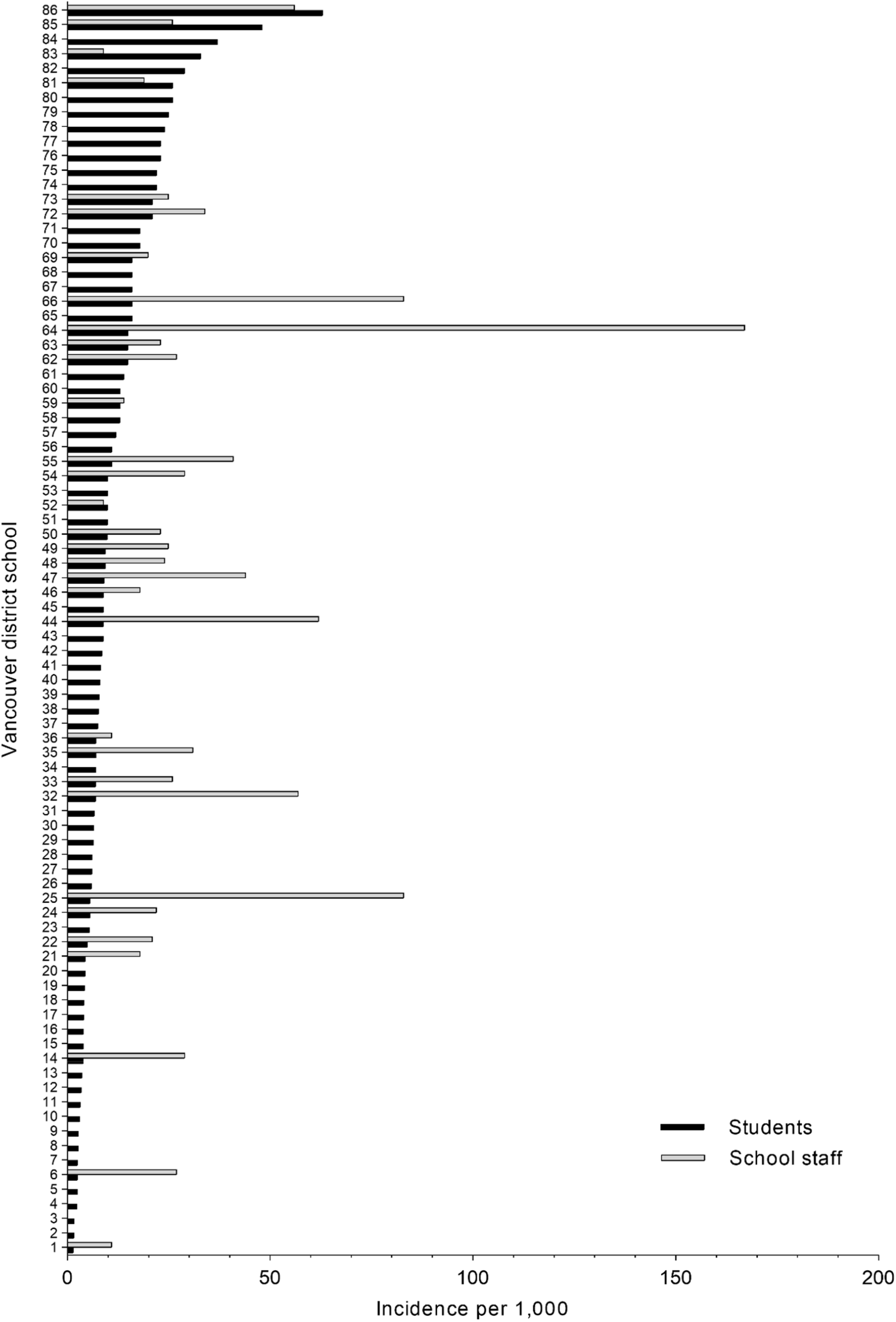
Incidence of reported COVID-19 cases (Vancouver Coastal Health) per students (n = 47,280) and staff (n = 7,071) between schools of the Vancouver School District. (21 of 107 schools had zero COVID-19 cases among students and staff, and are not included in this graph).

**Supplemental Table 1:** Reported positive viral testing among entire Vancouver School District staff.

| **Reporting period** | **Classroom staff\***<br>**N = 5091** | **Non-classroom staff\*\***<br>**N = 1408** | **Other school staff\*\*\***<br>**N = 572** | **Overall**<br>**N = 7071** |
| --- | --- | --- | --- | --- |
| Up to and including September 9<sup>th</sup>, 2020 | 7 | 1 | 1 | 9 |
| Between September 10<sup>th</sup> 2020 and March 4<sup>th</sup> 2021 inclusive (median date of completion of study questionnaires) | 58 | 18 | 8 | 84 |
| Between March 5<sup>th</sup> 2020 and April 23<sup>rd</sup> 2021 | 34 | 8 | 4 | 46 |

\*Teachers, Teacher Librarians, Resource Teachers, Student Support Workers, Family and Youth Workers, and Counsellors, including staff who are on call for these positions.
\*\*Principals/VPs, Office Administrative Assistants, Facilities Staff, Building Engineers, and Custodians, including staff who are on call for these positions.
\*\*\*Food services, Supervision Aides, and District staff, including staff who are on call for these positions.

**Supplemental Table 2:** Results of ORTHO (Spike-based) or ROCHE (N-based) serology testing, and according to self-reported viral PCR, for Spike-POSITIVE, VACCINATED cases (n=35)

| Antibody detection |  |  |  | Self-reported COVID-19 PCR result? | Days between vaccine and serology | Final case assignment |
| --- | --- | --- | --- | --- | --- | --- |
| Spike reactivity | Spike (index) | N reactivity | N (index) |  |  |  |
| R | 4.8 | NR | 0.082 | Not tested | 16 | Negative |
| R | 73.6 | NR | 0.089 | No | 14 | Negative |
| R | 252 | R | 150.7 | Yes | 1 | Positive |
| R | 142 | NR | 0.088 | No | 24 | Negative |
| R | 69.9 | R | 99.09 | Yes | 20 | Positive |
| R | 31.1 | NR | 0.084 | Not tested | 20 | Negative |
| R | 279 | NR | 0.144 | Not tested | 15 | Negative |
| R | 27.8 | NR | 0.09 | Not tested | 15 | Negative |
| R | 13.7 | NR | 0.088 | Not tested | 56 | Negative |
| R | 47.3 | NR | 0.087 | No | 32 | Negative |
| R | 163 | NR | 0.089 | Not reported | 11 | Negative |
| R | 10.5 | NR | 0.091 | No | 2 | Negative |
| R | 2.64 | NR | 0.09 | Not tested | 17 | Negative |
| R | 15.3 | NR | 0.092 | Not tested | 16 | Negative |
| R | 285 | NR | 0.082 | Not tested | 16 | Negative |
| R | 7.95 | NR | 0.085 | No | ? | Negative |
| R | 16.8 | NR | 0.086 | Not tested | 31 | Negative |
| R | 114 | NR | 0.087 | Not tested | 17 | Negative |
| R | 90 | NR | 0.082 | Not tested | 16 | Negative |
| R | 15.9 | NR | 0.096 | Not tested | 18 | Negative |
| R | 486 | R | 44.52 | Yes | 88 | Positive |
| R | 63.5 | NR | 0.09 | Not tested | ? | Negative |
| R | 1.07 | NR | 0.093 | No | 14 | Negative |
| R | 3.78 | NR | 0.095 | Not tested | 20 | Negative |
| R | 20.5 | NR | 0.212 | No | 21 | Negative |
| R | 132 | NR | 0.088 | No | 57 | Negative |
| R | 20.7 | NR | 0.095 | Not tested | 20 | Negative |
| R | 147 | NR | 0.085 | No | 21 | Negative |
| R | 61.5 | NR | 0.093 | Not tested | 24 | Negative |
| R | 106 | NR | 0.096 | Not tested | 23 | Negative |
| R | 73.6 | NR | 0.091 | Not tested | ? | Negative |
| R | 79.9 | NR | 0.094 | No | 35 | Negative |
| R | 2.47 | NR | 0.095 | Not tested | 16 | Negative |
| R | 9.71 | NR | 0.098 | No | 15 | Negative |
| R | 8.56 | NR | 0.094 | Not tested | 28 | Negative |

R: reactive; NR: non-reactive; the only 3 vaccinated staff that tested positive by both S- and N-based assays also reported a history of COVID-19 by positive PCR viral test, and none of the vaccinated school staff who tested negative by the N-based serology assay reported a positive PCR viral test. All spike (S)-reactive cases were contacted by phone or email to confirm the date they received a COVID-19 vaccine between the date of survey completion and blood sampling. Three participants indicated that they had received a COVID-19 vaccine, but omitted to indicate the date in the questionnaire or email response.

**Supplemental Table 3:** Results of ORTHO (Spike-based) or ROCHE (N-based) serology testing, and according to self-reported viral PCR, for seropositive, INFECTED cases (n=35)

| Antibody detection |  |  |  | Self-reported COVID-19 PCR result? | Time between PCR test and serology | Vaccinated? [days between vaccine and serology] |
| --- | --- | --- | --- | --- | --- | --- |
| Spike reactivity | Spike (index) | N reactivity | N (index) |  |  |  |
| R | 6.2 | R | 1.44 | Yes | 2 months | No |
| R | 224 | R | 57.99 | Not tested | - | No |
| R | 129 | R | 5.99 | Not tested | - | No |
| R | 112 | R | 127 | Yes | 1.9 month | No |
| R | 226 | R | 125.3 | Yes | 3.2 months | No |
| R | 433 | R | 64.64 | Not tested | - | No |
| R | 270 | R | 166.7 | Yes | 2.5 months | No |
| R | 8.73 | NR | 0.483 | Yes | 3 weeks | No |
| R | 272 | R | 6.41 | Yes | 3 months | No |
| R | 3.22 | NR | 0.074 | Not tested | - | No |
| NR | 0.39 | R | 8.61 | Yes | 11.25 months | No |
| R | 445 | R | 2.84 | No | - | No |
| R | 242 | R | 1.99 | Not tested | - | No |
| R | 52.4 | R | 4.58 | Yes | 5.5 months | No |
| R | 138 | NR | 0.564 | Not tested | - | No |
| R | 318 | R | 42.54 | Not tested | - | No |
| R | 2.27 | R | 117.6 | Yes | 2.75 months | No |
| R | 562 | R | 28.92 | Yes | ~7 months | No |
| R | 216 | R | 119.3 | Yes | 2.3 months | No |
| R | 381 | R | 193.3 | Not tested | - | No |
| R | 142 | R | 11.03 | Yes | 1.5 month | No |
| R | 331 | R | 3.54 | Not tested | - | No |
| R | 69.7 | R | 16.88 | Not tested | - | No |
| R | 252 | R | 150.7 | Yes | 2 months + 6 days | Yes [1] |
| R | 69.9 | R | 99.09 | Yes | 4.25 months | Yes [20] |
| R | 4.44 | R | 4.26 | Not tested | - | No |
| R | 45.1 | R | 4.22 | Not tested | - | No |
| R | 581 | R | 44.07 | Yes | 1 year + 2 weeks | No |
| R | 503 | R | 45.95 | Yes | 6.75 months | No |
| R | 10.3 | R | 1.01 | Yes | 2.75 months | No |
| R | 211 | R | 130.5 | Yes | 1.84 month | No |
| R | 310 | R | 80.05 | Yes | 4.5 months | No |
| R | 224 | R | 15.64 | Not tested | - | No |
| R | 327 | R | 57.04 | Yes | 4 months + 2 days | No |
| R | 486 | R | 44.52 | Yes | 3.75 months | Yes [8] |

R: reactive; NR: non-reactive; One school staff who tested positive for COVID-19 by PCR viral test tested negative by serology ~5 months later both by S- and N-based assays. Another who reported a positive PCR test did not complete the serology testing.

## SUPPLEMENTAL DATA: Matching scheme of school workers with Canadian blood donors

1537 observations were matched from school workers, with 24 missing postcode, 6 missing sex.

In January, seroprevalence data were available from 4910 adults in the lower mainland of British Columbia west of Kamloops and Kelowna.

A multiple step procedure aiming for 1:2 matching was employed as shown below.

|  |  |  | Seropositive Canadian blood donors N (%) |  |
| --- | --- | --- | --- | --- |
| Matching criteria | Number of school staff with 2 matched donors found | Number of matched blood donors | Roche N, N (%) | Roche S, N (%) |
| Age ± 2 years, same sex, and first two digits of postal code | 671 | 1342 | | |
| Age ± 2 years and same sex | 429 | 858 | | |
| Age ± 5 years and same sex | 18 | 36 | | |
| Age ± 10 years and same sex | 63 | 126 | | |
| Age ± 15 years and same sex | 70 | 140 | | |
| Total | 1251 | 2502 | 51<br>(2.03%) | 51*<br>(2.03%) |

\*23 vaccinated, S-based assay positive, N-based assay-negative were moved to negative group.

## APPENDIX 1 COVID-19 mitigations measures in Vancouver schools (2020/21 school year)

Prior to reopening, the District implemented COVID-19 safety plans consistent with the British Columbia Centre for Disease Control (BCCDC) COVID-19 Public Health Guidance for K-12 School Settings: http://www.bccdc.ca/Health-Info-Site/Documents/COVID_public_guidance/Guidance-k-12-schools.pdf. and Provincial COVID-19 Health & Safety Guidelines for K-12 Settings: https://www2.gov.bc.ca/assets/gov/education/administration/kindergarten-to-grade-12/safe-caring-orderly/k-12-covid-19-health-safety-guidlines.pdf with support from Public Health.

COVID-19 safety plans included public health measures (e.g., protocols for testing and contact tracing), environmental measures (e.g., maximization of distance in classrooms, enhanced cleaning and disinfection, improved fresh air intake), administrative measures (e.g., staggered scheduling, assigning students and staff cohorts), personal measures (e.g., daily symptom checks, physical distancing, hand hygiene, respiratory etiquette), and personal protective equipment.

At the beginning of the school year (late August/September), parents were given the option of: 1) full time in-person schooling, 2) home schooling, 3) online learning and 4) a temporary online learning with return to full time schooling with re-entry dates offered later in fall and in January 2021.

Daily health assessments were required by all staff and students (via parents) prior to arriving at school and again upon arrival. Anyone with even minor symptoms of cold or flu-like illness was to stay home or go home if these symptoms developed mid-day. Classrooms and other spaces were arranged to maximize distance between students and staff. Class sizes were set by grades: 20 students / class for kindergarten; 22 for grades 1 to 3; 30 for grades 4 to 12. School staff and their students were assigned specific classrooms which were between 75 m – 83 m for elementary students (K to grade 7) and 75 m – 80 m for secondary students (grades 8 to 12) with larger spaces available for elective courses (e.g., physical education, food studies, metal, woodworking, automotive). In addition, secondary classrooms were divided into two separate groupings (AM and PM) of 15 students.

The plan also included revising school schedules and learning groups where students also had staggered recess and lunch breaks, and were assigned specific outside areas. Elementary students (K-7) received full day in-class instruction in their assigned learning groups/cohorts. Secondary students (grades 8 to 12) had both in-class instruction and remote learning and their schedules shifted to a quarter system with maximum two in-person classes a day with further instruction given remotely.

Ventilation measures included opening windows to promote fresh air flow to classrooms as well as indoor air ventilation improved with the HVAC systems running longer during the day, recirculating less air and the filters changed to higher efficiency (MERV13) filters.

Other measures included, the addition of hand sanitizer to classrooms and common areas, directional traffic flow within the school, provision of plexiglass as needed for certain staff roles, and the training of all staff on the safety plan and protocols. In addition to the regular daily cleaning by custodial staff, twice daily disinfection of all high touch frequency items was conducted. Shared items in classrooms were limited and the teachers, or if in secondary school, the students disinfected those used. Initially, masking was encouraged, but not required. Non-medical mask use was encouraged but not required early in the school year, and then in February 2021 masks were required for all staff and students in grades 6-12 in common spaces, and ultimately from April onward for students grades 4-12 at all times while indoors at school. This guidance did not apply if staff or students did not tolerate a mask for health or behavioural reasons. Masks remained recommended for K-3 students. Two reusable cloth masks were distributed to all staff and students in September 2020 and again in January 2021.

SARS-CoV-2 nucleic acid amplification (PCR) was available for anyone with symptoms through the provincial health system, and advised for students or staff with fever or new symptoms which persisted for over 24 hours. Tests were generally processed within 24 hours, and positive tests were automatically reported to Public Health which investigated cases within 24 hours, and initiated contact tracing.

Symptomatic close contacts were asked to seek testing; asymptomatic testing was not used to release contacts from isolation on an earlier timeline. All close contacts, including close contacts at school, were isolated for at least 14 full days. Entire classes were not isolated unless all members were identified as close contacts. School closures to control transmission were not required during the study period. Also, at the time, vaccination programs had not substantially reached working-age people until April 14, and the majority of school staff vaccinations occurred in May.

